# Apnea, Intermittent Hypoxemia, and Bradycardia Events Predict Late-Onset Sepsis in Extremely Preterm Infants

**DOI:** 10.1101/2024.01.26.24301820

**Authors:** Sherry L. Kausch, Douglas E. Lake, Juliann M. Di Fiore, Debra E. Weese-Mayer, Nelson Claure, Namasivayam Ambalavanan, Zachary A. Vesoulis, Karen D. Fairchild, Phyllis A. Dennery, Anna Maria Hibbs, Richard J. Martin, Premananda Indic, Colm P. Travers, Eduardo Bancalari, Aaron Hamvas, James S. Kemp, John L. Carroll, J. Randall Moorman, Brynne A. Sullivan, The Prematurity-Related Ventilatory Control (Pre-Vent) Investigators

## Abstract

**Objectives:** Detection of changes in cardiorespiratory events, including apnea, periodic breathing, intermittent hypoxemia (IH), and bradycardia, may facilitate earlier detection of sepsis. Our objective was to examine the association of cardiorespiratory events with late-onset sepsis for extremely preterm infants (<29 weeks’ gestational age (GA)) on versus off invasive mechanical ventilation.

**Study Design:** Retrospective analysis of data from infants enrolled in Pre-Vent (ClinicalTrials.gov identifier NCT03174301), an observational study in five level IV neonatal intensive care units. Clinical data were analyzed for 737 infants (mean GA 26.4w, SD 1.71). Monitoring data were available and analyzed for 719 infants (47,512 patient-days), of whom 109 had 123 sepsis events. Using continuous monitoring data, we quantified apnea, periodic breathing, bradycardia, and IH. We analyzed the relationships between these daily measures and late-onset sepsis (positive blood culture >72h after birth and ≥ 5d antibiotics).

**Results:** For infants not on a ventilator, apnea, periodic breathing, and bradycardia increased before sepsis diagnosis. During times on a ventilator, increased sepsis risk was associated with longer IH80 events and more bradycardia events before sepsis. IH events were associated with higher sepsis risk, but did not dynamically increase before sepsis, regardless of ventilator status. A multivariable model predicted sepsis with an AUC of 0.783.

**Conclusion:** We identified cardiorespiratory signatures of late-onset sepsis. Longer IH events were associated with increased sepsis risk but did not change temporally near diagnosis. Increases in bradycardia, apnea, and periodic breathing preceded the clinical diagnosis of sepsis.

## Introduction

Premature infants require prolonged hospitalization in the Neonatal Intensive Care Unit (NICU), during which time they undergo continuous cardiorespiratory monitoring. Up to 15% of preterm infants with GA< 29 weeks develop late-onset sepsis, a potentially severe or even fatal bloodstream infection (1–3). The systemic inflammatory response to infection causes changes in the control of heart rate and breathing, regulated by the autonomic nervous system (4–6). Impaired autonomic control during sepsis can manifest as apnea, at least in part due to release of endogenous prostaglandins (7, 8). In premature infants, apnea episodes are often accompanied by bradycardia. Apnea and intermittent hypoxemia (IH) frequently occur during the NICU course even in the absence of sepsis, as a result of immature control of breathing and lung disease (9), and transient bradycardia events may occur with vagus nerve activation with or without sepsis (5, 10, 11). Periodic breathing, repetitive cycles of short apneic pauses interspersed with tachypneic periods, is another common physiologic phenomenon in preterm infants that may increase in the setting of sepsis (9, 12, 13).

Earlier detection and treatment of sepsis, prior to overt clinical deterioration, can improve outcomes. One way to do this is through continuous analysis of cardiorespiratory data to detect subtle signatures of illness not apparent during routine monitoring. One example is heart rate (HR) characteristics monitoring, with application of algorithms to detect reduced HR variability and increased transient HR decelerations to predict clinical deterioration with a sepsis-like illness in the subsequent 24-hours (14–16). Adding analysis of pulse oximetry-derived oxygen saturation (SpO_2_) and cross-correlation of HR and SpO_2_ (which can detect concurrent bradycardia-desaturation events due to apnea) can further inform on risk of sepsis (17–19).

Large, multicenter datasets allow for development and validation of generalizable models of late-onset sepsis. For the current work, we analyzed data archived by the Prematurity-Related Ventilatory Control (Pre-Vent) consortium. The main objective of Pre-Vent was to study the frequency and severity of apnea, bradycardia, IH, and periodic breathing throughout the NICU course of extremely preterm infants at five level IV NICUs to understand how these events relate to adverse respiratory outcomes (20, 21). The algorithms developed from this work capture events that clinicians in the NICU recognize at the bedside but cannot usually quantify in an automated manner. Our aims in the current work were to use data from Pre-Vent to: (A) examine the association of cardiorespiratory metrics with sepsis while accounting for the effect of invasive mechanical ventilation, (B) assess the dynamic changes in cardiorespiratory metrics preceding sepsis, and (C) develop multivariable models incorporating cardiorespiratory signatures to predict the diagnosis of blood culture-positive late-onset sepsis.

## Methods

### Study design and patient population

We used data from an observational study of infants born <29 weeks’ gestational age (GA) prospectively enrolled in Pre-Vent (ClinicalTrials.gov identifier NCT03174301).(21) Institutional Review Boards (IRBs) reviewed the study and approved collection of clinical and cardiorespiratory monitoring data at all participating centers with a waiver of consent at three centers and informed consent required at two centers. Oversight was also provided by an observational and safety monitoring board, appointed by the NHLBI. Additional details are included in an **online data supplement.** This study followed the Transparent Reporting of a Multivariable Prediction Model for Individual Prognosis or Diagnosis (TRIPOD) checklist: Prediction Model Development and Validation.

### Clinical data collection and definitions

Clinical data were recorded in a REDCap database and consisted of demographic and clinical variables, including late-onset sepsis events and daily recordings of respiratory support. We defined late-onset sepsis as a positive blood culture obtained after 72 hours of age and treated with ≥5 days of antibiotics or a shorter duration if the infant died during treatment. A clinician at each site reviewed blood cultures to confirm the diagnosis of sepsis. Multiple sepsis events for the same infant were included if separated by at least 10 days. Respiratory support was dichotomized at daily intervals as either on or not on invasive mechanical ventilation with an endotracheal tube. The day of intubation was recorded as a ventilator day and the day of extubation was recorded as a non-ventilator day. Days not on invasive mechanical ventilation included days on room air or non-invasive respiratory support devices such as nasal cannula, continuous positive airway pressure, or non-invasive positive pressure ventilation.

### Cardiorespiratory metrics and definitions

Cardiorespiratory metrics were calculated from bedside monitor waveform and vital sign data using algorithms and criteria described in the primary Pre-Vent study (21). The metrics were quantified as follows:

1. Apnea: average daily counts of pauses in breathing for ≥ 20 seconds derived from chest impedance waveforms (22)
2. Periodic breathing: minutes per day in periodic breathing, calculated using a wavelet transform and a 5-breath template from chest impedance waveforms (12)
3. IH80 DPE: average duration of events with SpO_2_ < 80% for 10 to 300 seconds (23)
4. IH90 DPE: average duration of events with SpO_2_ < 90% for 10 to 300 seconds (23)
5. Bradycardia: average daily counts of a drop in HR to <80 bpm for at least 5 seconds (21)

Because mechanical ventilation modifies respiratory efforts, we did not quantify apnea or periodic breathing during times on a ventilator. IH and bradycardia metrics were calculated both on and off the ventilator. Additional details are reported in the **online data supplement** (supplement page 3).

For univariate modeling, we included all available daily measures for each of the five features irrespective of missing vital sign or waveform data. For multivariable modeling, we excluded days without physiologic monitoring data available in order to be able to calculate all five cardiorespiratory metrics.

### Analytic Approach

We used Wilcoxon rank sum tests and chi-squared tests to compare continuous and categorical clinical variables among infants with and without one or more episodes of sepsis and to compare variables during periods on and off ventilator.

Univariate and multivariable logistic regression models were developed using daily values of cardiorespiratory features and clinical variables as predictors and sepsis within the subsequent three days as the outcome. The day of the positive blood culture and the two days preceding it were classified as sepsis. The ten days following sepsis were excluded from analysis to remove periods of sepsis treatment and recovery. All other time periods were labeled as no sepsis. We evaluated the relationships between cardiorespiratory features and clinical variables with sepsis using univariate logistic regression models for times off and on a ventilator. We used plots of predicted sepsis risk to visualize ranges of clinical and cardiorespiratory feature values associated with significantly increased or decreased risk of sepsis, depicted by red line segments to indicate where the 95% confidence interval around the predicted risk of sepsis did not cross one (the average risk of sepsis).

To evaluate for dynamic changes in cardiorespiratory features around the time of sepsis, we made time-to-event plots, grouped by ventilator status, displaying the 10-day period around sepsis diagnosis. To determine if a cardiorespiratory feature was increasing in the three days leading up to sepsis, we tested the null hypothesis that cardiorespiratory feature values are less than or equal to the value for the same patient 1 day prior using Wilcoxon rank sum test.

To evaluate the combined ability of cardiorespiratory and clinical features to predict sepsis, we used multivariable logistic regression with varying numbers of features. Internal validation of the multivariable logistic regression models was evaluated using 10-fold cross-validation to obtain the area under the receiver operator characteristic curve (AUC). Goodness-of-fit was assessed by log loss and normalized to values between zero and one using McFadden’s pseudo r-squared (25). We used goodness-of-fit to assess how well the model fits the data. Calibration of the final model was assessed using a calibration plot. Feature importance was analyzed by comparing the decrease in cross-validated AUC loss introduced by removing that feature from the final multivariable model. Modeling was performed in R 4.2.3 (R Foundation for Statistical Computing, Vienna, Austria) using the *rms* package (26).

To account for repeated daily measures, including repeat sepsis events, we corrected for unequal variances and correlated measures from individual patients and sibling clusters. The statistical significance of the logistic regression model coefficients were adjusted for repeated measures using the Huber-White method to modify the variance-covariance matrix in all univariate and multivariable logistic regression models (27).

## Results

### Cohort characteristics

We analyzed clinical data from 737 infants enrolled in the Pre-Vent study with mean GA 26.4 weeks (SD = 1.7) (Table 1, Table S1). Cardiorespiratory monitoring data were missing from 18 (2%) of infants. Thus, cardiorespiratory analyses and multivariable modeling included 719 infants. Infants diagnosed with one or more episodes of late-onset sepsis had lower gestational age and birth weight than those without and were more likely to be small for gestational age while other demographic and perinatal variables analyzed were similar (**Table 1, Figure S1**). Causative organisms were Coagulase-negative staphylococci (36%), *Staphylococcus aureus* (15%), *E. coli* (10%), other Gram-positives (7%), and other Gram-negatives (32%). In total, we analyzed 47,512 patient-days with monitoring data available (79% of NICU days for the 719 infants), of which 12,634 (26%) were ventilator days (**Table 2**). For univariate analysis, we used all times data were available to calculate each of the five features (see **Table S2**). Sepsis occurred more often during times on a ventilator, and sepsis on a ventilator occurred at a lower postmenstrual age and postnatal age than sepsis not on a ventilator (**Table 2**).

**Table 1.** Perinatal and demographic information by outcome.

| Characteristic | Combined Population (N=737) | Infants with Sepsis (N=133) | Infants without Sepsis (N=604) | p-value<sup>a</sup> |
| --- | --- | --- | --- | --- |
| Birth weight (grams) (mean ± SD) | 870 ± 260 | 746 ± 226 | 897 ± 260 | <0.001 |
| Gestational age (weeks) (mean ± SD) | 26.4 ± 1.7 | 25.5 ± 1.7 | 26.6 ± 1.6 | <0.001 |
| Small for gestational age | 86 (11.7%) | 24 (18.0%) | 62 (10.3%) | 0.017 |
| Race |  |  |  |  |
| Black | 357 (48.4%) | 67 (50.4%) | 290 (48.0%) | 0.691 |
| White | 324 (44.0%) | 58 (43.6%) | 266 (44.0%) | 0.562 |
| Other | 56 (7.6%) | 8 (6.0%) | 48 (7.9%) | 1.000 |
| Male sex | 375 (50.9%) | 67 (50.4%) | 308 (51.0) | 0.974 |
| Outborn birth | 65 (8.8%) | 13 (9.8%) | 52 (8.6%) | 0.795 |
| Mode of birth: C-section | 499 (67.7%) | 84 (63.2%) | 415 (68.7%) | 0.256 |

Values are either mean ± standard deviation or number and percentage. <sup>a</sup> P-values refer to comparisons between infants with and without late-onset sepsis were calculated using Wilcoxon rank sum tests for continuous variables or chi-square goodness of fit test for proportions. SD: standard deviation. PNA: postnatal age

**Table 2.**
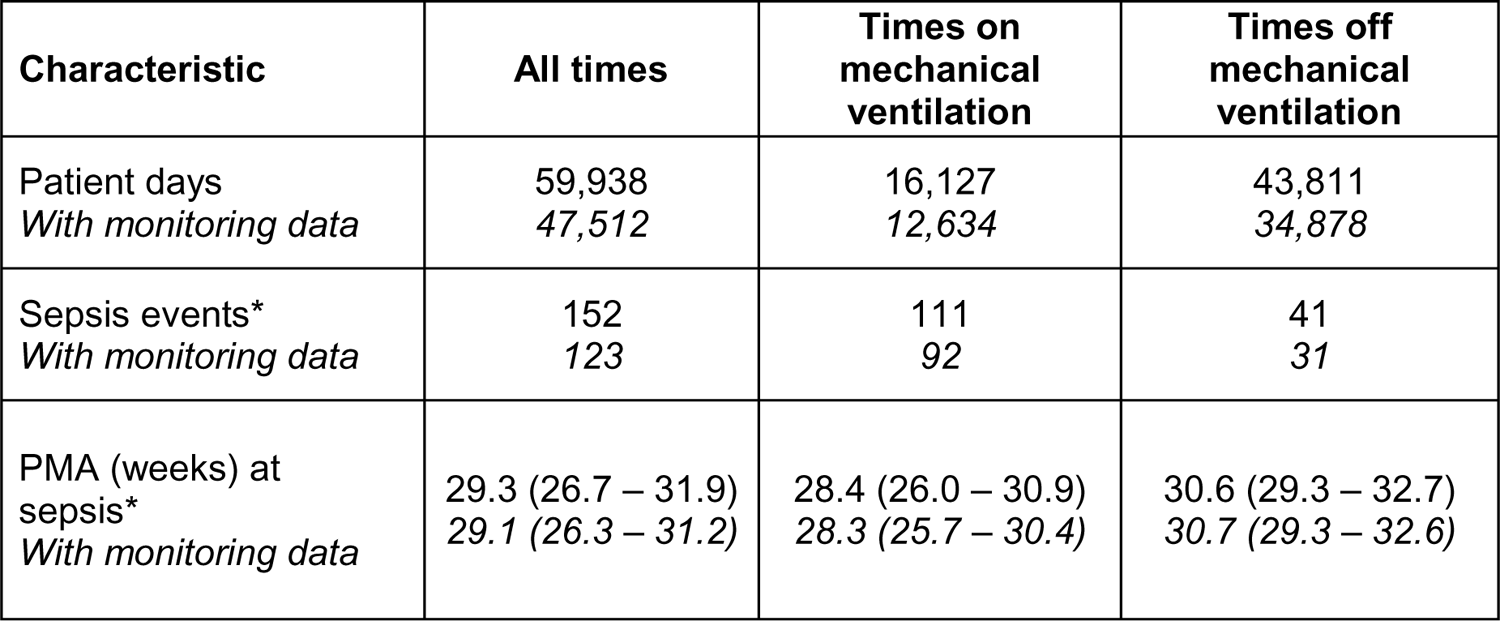
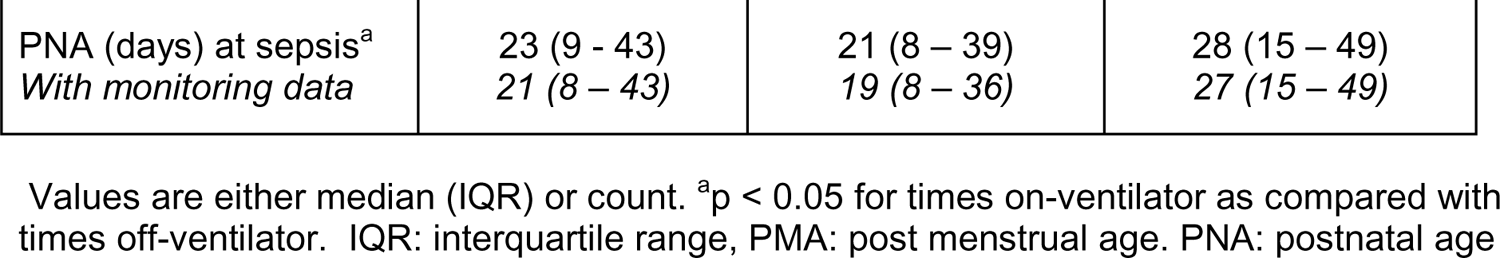
Characteristics of the study population grouped by data collected during all times and during periods on and off invasive mechanical ventilation.

### Univariate analysis results

Figure 1 and **2** depict univariate modeling results as the relative risk of sepsis diagnosis in the subsequent three days for clinical variables and cardiorespiratory metrics, respectively, during times off and on a ventilator. Figure 3 shows the average value of cardiorespiratory features over five days before and after the clinical diagnosis of sepsis, stratified by ventilator status. These figures allowed us to visualize results to make qualitative as well as quantitative interpretations of individual clinical and cardiorespiratory predictors.

**Figure 1.**
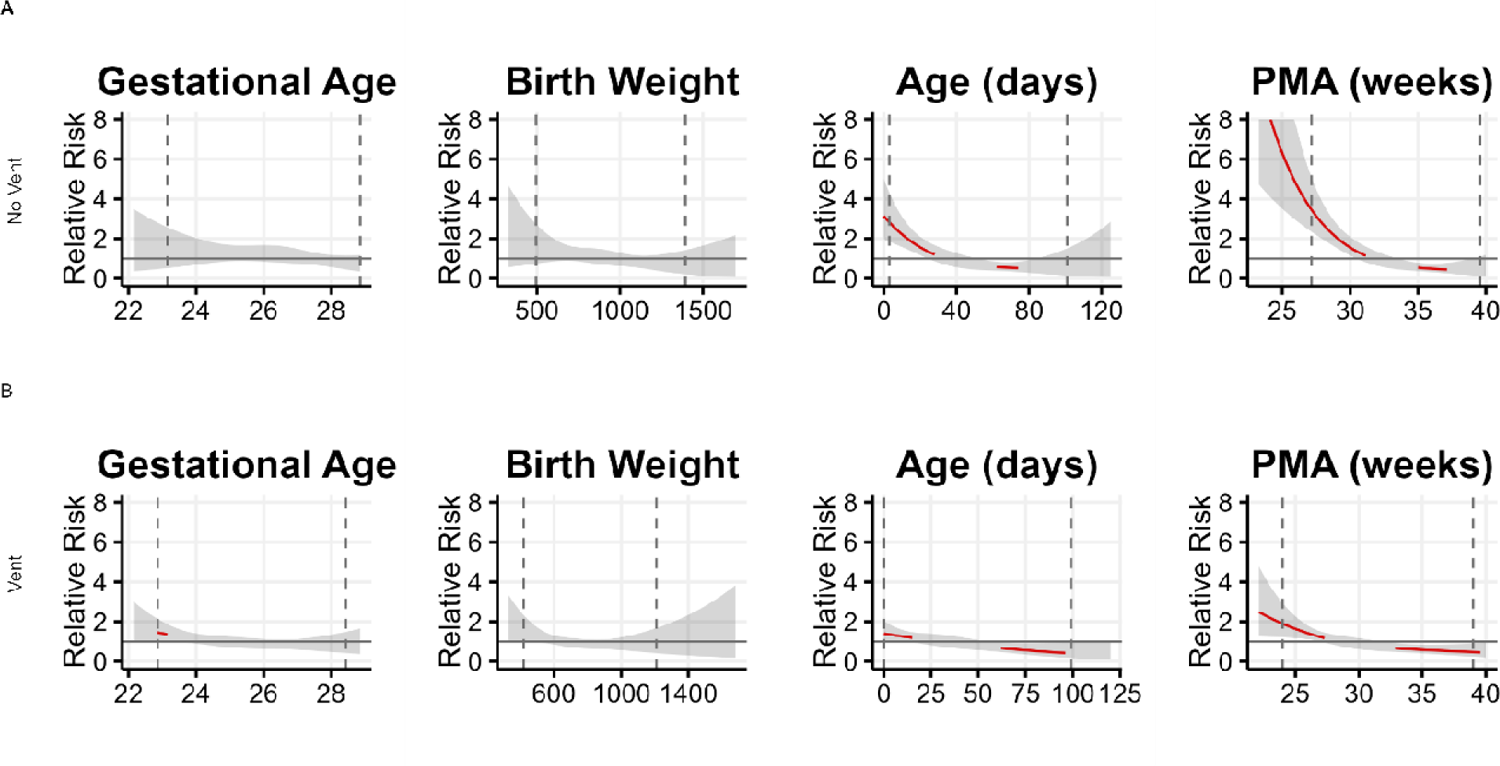
Sepsis risk based on clinical predictors. Depiction of associations of predictors in each univariate logistic regression model for times (A) off-ventilator (No Vent) and (B) on-ventilator (Vent). Each tile is a plot of the relative risk of sepsis as a function of the predictor across its range, where 1 on the y-axis indicates risk equal to the overall sepsis event rate. The event rate for on-ventilator times was 0.0188 and for off-ventilator times was 0.0036. The translucent ribbon represents the 95% confidence interval. The vertical dashed lines indicate the 2.5 and 97.5 percentile of data. The red line highlights the range where the confidence intervals do not include 1. In these ranges, the variable can be considered a predictor of significantly increased sepsis risk. The major finding is increased sepsis risk at lower PMA.

**Figure 2.**
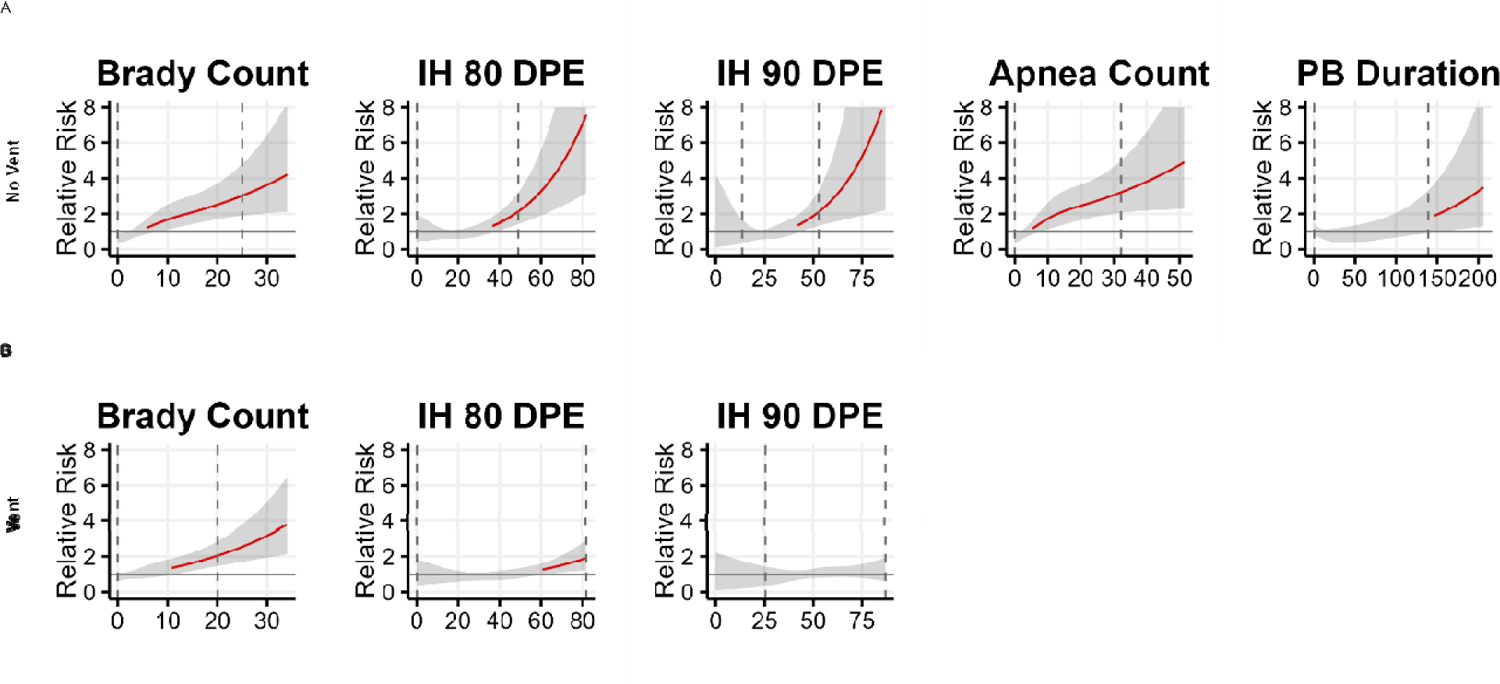
Sepsis risk based on cardiorespiratory predictors, on and off invasive mechanical ventilation. Associations of predictors in each univariate logistic regression model for times (A) off-ventilator and (B) on-ventilator. Brady count is the number of bradycardia events per day; IH is intermittent hypoxemia; DPE is duration per event in seconds; PB is periodic breathing exposure in minutes per day. The rate of sepsis per day for times on-ventilator are 0.0188 and for times off-ventilator 0.0036. The major findings are increased sepsis risk when cardiorespiratory measures are increased.

**Figure 3.**
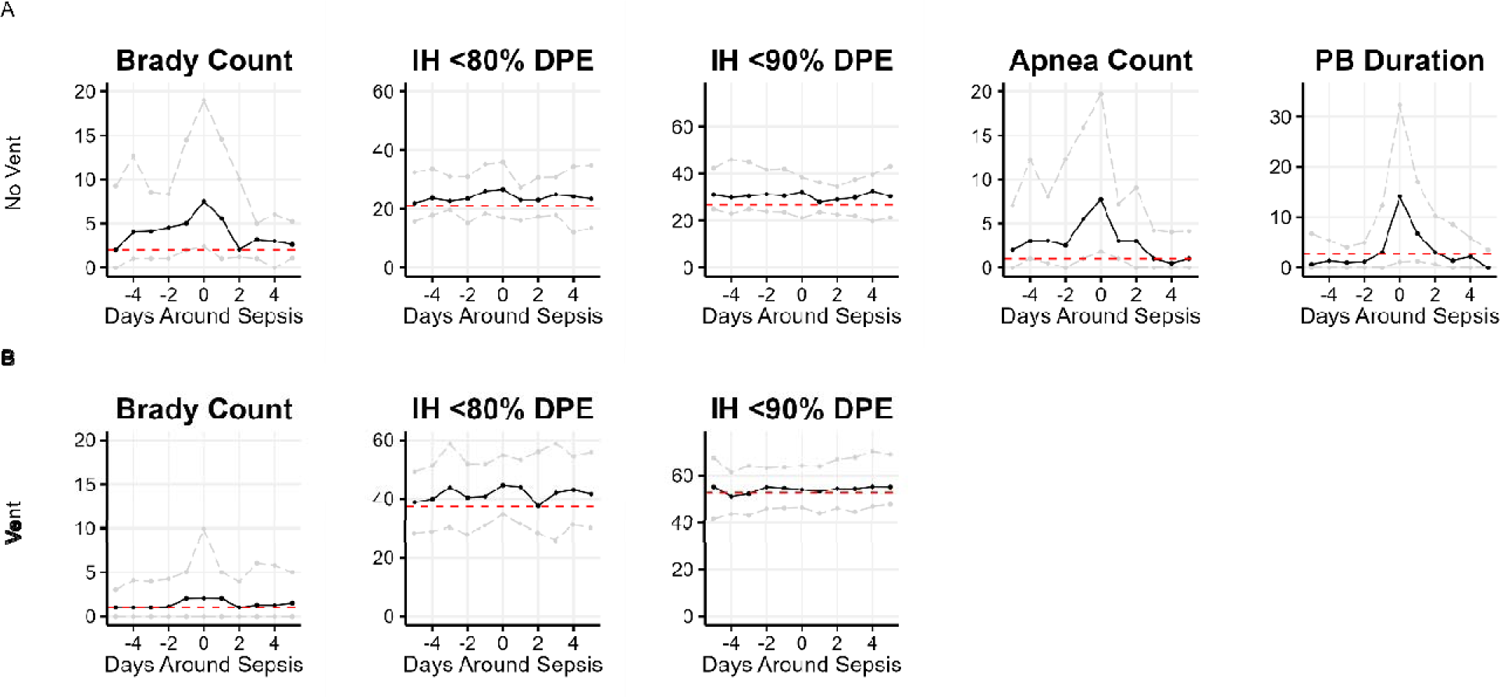
Cardiorespiratory events around sepsis diagnosis, on and off invasive mechanical ventilation. Panels show the time courses of cardiorespiratory events 5 days before and after diagnosis of sepsis at day zero (A) off-ventilator and (B) on-ventilator. Solid lines represent the median value; gray dashed lines represent the 25th and 75th percentiles; horizontal red lines represent the median value for all times irrespective of sepsis. Brady count is the number of bradycardia events per day; IH is intermittent hypoxemia; DPE is duration per event in seconds; PB is periodic breathing exposure in minutes per day. The major finding is of dynamic changes in bradycardia, apnea and periodic breathing but not in IH events. Daily median bradycardia counts and the median IH80 and IH90 DPE are shown for times on- and off-ventilator (A, B). Additionally, for times off-ventilator (A), median daily number of apnea events and median duration per event for PB are shown.

Of the clinical variables, PMA was the strongest predictor, with lower post menstrual age associated with significantly increased sepsis risk and higher postmenstrual age associated with significantly decreased sepsis risk. (Figure 1, **Table S1**). Higher levels of respiratory support and FiO_2_ were also associated with increased sepsis risk (**Table S3**). For times not on a ventilator, increased sepsis risk was associated with more bradycardia and apnea events, longer duration of IH80 and IH90 events, and more time in periodic breathing (Figure 2A, red lines). During times on a ventilator, increased risk of sepsis was associated with longer duration of IH80 events, and higher number of bradycardia events (Figure 2B, red lines).

Next, we plotted features as a function of the time to sepsis and performed sign rank tests to identify significant changes in measures from the same patient one day prior (Figure 3). For infants not on a ventilator, the frequency of apnea and bradycardia events and the exposure to periodic breathing per day increased in the 1-2 days before sepsis diagnosis (Figure 3A).

While higher duration of IH80 events was a sepsis risk marker for infants not on a ventilator, the duration of IH events did not increase significantly near the time of sepsis diagnosis.

For infants on a ventilator, the duration of IH80 events significantly increased from the day before to the day of sepsis diagnosis. (Figure 3B, p<0.05 comparing time −1 day to time 0). Visually, the median duration of IH events did not have a clinically meaningful increase in any of the 3 days prior to sepsis (Figure 3B).

### Multivariable modeling results

We developed multivariable predictive models for the diagnosis of late-onset sepsis in a stepwise fashion to evaluate the added information gained from different variables. While PMA alone resulted in an AUC 0.739, adding cardiorespiratory variables available at all times irrespective of ventilator status (IH80 and bradycardia count) increased the AUC to 0.763.

Adding ventilator status and cardiorespiratory variables only available during times not on a ventilator (apnea count and periodic breathing exposure) resulted in a model with an AUC of 0.783. Model fit was improved with the addition of cardiorespiratory variables. **Table 3** summarizes multivariable models and results. The model with all features had good calibration (Figure S2). IH90 events had little impact on models, and were excluded.

**Table 3.** Area under the receiver operating characteristic curve for multivariable models when progressively more features are included.

| Model | Variables | cvAUC | McFadden's pseudo R<sup>2</sup> |
| --- | --- | --- | --- |
| Clinical Variables Alone | PMA | 0.739 | 0.067 |
| Clinical Model + 2 Physiologic Variables | PMA + IH 80 + Bradycardia | 0.763 | 0.081 |
| Clinical Model + 2 Physiologic Variables + Ventilator status | PMA + IH 80 + Bradycardia + Vent status | 0.782 | 0.092 |
| Clinical Model + 4 Physiologic Variables + Ventilator status | PMA + IH 80 + Bradycardia + Vent status + Apnea + Periodic Breathing | 0.783 | 0.094 |

Models with progressively more features are evaluated using 10-fold cross validation for AUC and McFadden's pseudo R<sup>2</sup> to assess model fit. AUC: Area under the receiver operating characteristic curve

We tested models for times on and off a ventilator separately and combined using bivariate logistic regression. This approach did not improve performance over the final multivariable model described in **Table 3**.

## Discussion

In this secondary analysis of continuous bedside monitor data from extremely premature infants archived by the Pre-Vent consortium at 5 NICUs, we studied the association of common cardiorespiratory events with the diagnosis of late-onset sepsis. We found that quantitative measures of IH, bradycardia, apnea, and periodic breathing predict sepsis and add to PMA, which was the strongest risk factor. These cardiorespiratory sepsis risk markers had either static or dynamic trends in frequency or duration near clinical diagnosis. Mechanical ventilation was an important variable to consider when using cardiorespiratory features to predict late-onset sepsis.

As with all adverse events in preterm infants, late-onset sepsis is associated with lower gestational age and PMA, and we found that PMA was superior to other demographic variables for sepsis prediction. We found that adding measures of cardiorespiratory instability provided incremental improvement in multivariable modeling, with cross-validated AUCs ranging from 0.739 to 0.783. In this work we used average daily summary metrics of events but note that these performance metrics compare with similar models developed in our other work in which we deployed HR and SpO_2_ metrics calculated at more frequent intervals (18, 19).

For infants not on invasive mechanical ventilation, transient bradycardia events increased in the day preceding sepsis diagnosis, a finding that aligns with our prior research. More than twenty years ago, in developing a heart rate characteristics index (HeRO score) as a sepsis predictor, reduced variability of HR punctuated by transient decelerations of even mild degree was a signature of impending sepsis in preterm infants (28, 29). Reduced HR variability and decelerations also occur in distressed fetuses in the setting of uteroplacental insufficiency or chorioamnionitis (30, 31). Bradycardia events in preterm infants may occur in conjunction with apnea (24), but in the current work we found them to be associated with imminent sepsis both off and on mechanical ventilation. In preclinical models we have found that other physiologic mechanisms of HR decelerations during sepsis aside from apnea include release of pro-inflammatory cytokines (32, 33) and activation of the vagus nerve (5).

For infants not on a ventilator at the time of sepsis, there was an increase in apnea and periodic breathing around the time of illness. Prostaglandins produced as part of the host response to infection depress respiration (7, 8) and pro-inflammatory cytokines impact control of breathing at the level of the brainstem in preclinical models (4). We have found in prior(19) and current work that cardiorespiratory algorithms to predict sepsis perform better for infants not on mechanical ventilation, which stands to reason since apnea is a major sign of illness and masked by the set breathing rate on a ventilator.

The average duration of hypoxemia events predicted sepsis both on and off a ventilator, but with a static risk relationship. In other words, prolonged IH at any time was associated with a higher risk of developing sepsis, in contrast to dynamic risk markers that increase in frequency or severity near the time of diagnosis. The mechanism for this risk relationship may be driven by the detrimental effects of IH on intestinal barrier function and microbiome (34–36), allowing for translocation of bacteria into the bloodstream. Alternatively, immaturity could be the driving risk factor of the association of prolonged IH with sepsis. IH events are universal in extremely preterm infants, reflecting both lung disease and immature control of breathing. We chose to focus on duration rather than frequency of IH events because the durations per event better predicted unfavorable respiratory outcomes in the primary analysis of the Pre-Vent cohort (21) and in other work in a different cohort were optimal for predicting death or BPD (37). The major strength of this work is access to a large archive of bedside monitor waveform and HR and SpO_2_ data from extremely preterm infants at multiple NICUs. Using these data, we were able to characterize static and dynamic changes in cardiorespiratory events near sepsis.

Understanding these risk relationships can help clinicians identify infants at high risk. When we can determine that signatures of illness are present, we can contemplate using statistical models as bedside predictive tools. Also, the findings in this cohort align with our prior studies of cardiorespiratory metrics predicting sepsis in very low birthweight infants at several other NICUs (18, 19). Previously, we used continuous calculations of the mean, standard deviation, skewness, kurtosis, and cross-correlation of HR and SpO_2_ to characterize signatures of illness due to late onset sepsis. The features used to predict sepsis in this study differ from our prior work in that apnea, bradycardia, and IH are intermittent events easily recognized by clinicians as being relevant to sepsis.

We note limitations. First, we did not account for all clinical variables that impact HR and SpO_2_ patterns of extremely preterm infants including changes in mode and settings of respiratory support. Second, the cardiorespiratory features used in our models were developed using pre-specified thresholds selected based on the results of the parent study and it is possible that alternative thresholds could improve the model performance. Finally, the data were analyzed at the daily level, which could miss changes in cardiorespiratory markers occurring with a rapid evolution of sepsis.

We measured markers of cardiorespiratory events in a quantitative manner using high-resolution bedside monitoring data. Our results characterize associations between apnea, bradycardia, and desaturation events and sepsis using a large, multicenter study. Our models were intended to test hypotheses rather than be implemented at the bedside to prompt evaluation. The information gleaned from this work add to our knowledge of physiology and sepsis in preterm infants <29 weeks GA.

## Conclusion

In a large multicenter cohort of extremely preterm infants with recording of continuous waveform and vital sign data, we identified cardiorespiratory metrics that predicted sepsis with both static and dynamic trajectories near the time of the positive blood culture. Increases in apnea, periodic breathing, and bradycardia events precede the diagnosis of sepsis. Models incorporating static and dynamic cardiorespiratory signatures and baseline risk factors could improve timely diagnosis and thus prognosis of preterm infants with late onset sepsis.

## List of Abbreviations

IH: (intermittent hypoxemia)

IH80: (intermittent hypoxemia with the average duration of events with SpO_2_ < 80% for 10 to 300 seconds)

IH90: (intermittent hypoxemia with the average duration of events with SpO_2_ < 90% for 10 to 300 seconds)

GA: (gestation age)

SD: (standard deviation) HR (heart rate)

NICU: (neonatal intensive care unit)

SpO_2_: _(_pulse oximetry-derived oxygen saturation)

Pre-Vent: (Prematurity-Related Ventilatory Control consortium)

AUC: (area under the receiver operator characteristic curve)

MAP: (mean airway pressure)

FiO_2_: (fraction of inspired oxygen) PMA (post menstrual age)

## Data Availability

All data produced in the present study are available upon reasonable request to the authors.

## Acknowledgements

The National Institutes of Health and the National Heart, Lung, and Blood Institute (NHLBI) provided grant support through cooperative agreements. While NHLBI staff did have input into the study design, conduct, analysis, and manuscript drafting, the content and views expressed are solely the responsibility of the authors and do not necessarily represent the official views of the National Institutes of Health or the U.S. Department of Health and Human Services.

Participating sites collected and stored the data while the University of Virginia, the lead data coordinating center (LDCC), analyzed the data. The co-PIs at each site had full access to his/her individual site data and take responsibility for the integrity of the raw waveforms while Drs. Randall Moorman (LDCC co-PI) and Douglas Lake (LDCC co-PI) take responsibility for the integrity of the data and accuracy of the data analysis.

We are indebted to our medical and nursing colleagues and the infants and their parents who agreed to take part in this study. Details of individual Pre-Vent Investigators are listed in the supplemental appendix.

## Author Contributions

SLK, KDF, and BAS conceptualized the study, planned the methodology, wrote the first draft of the manuscript, and reviewed and edited all subsequent drafts. SLK and DEL performed or supervised data curation, formal analysis, and prepared data visualizations. JMD, DEW-M, NC, NA, ZAV, KDF, CPT, and JRM reviewed the methodology and results and made substantial edits to the manuscript. PAD, JRM, DEL, AMH, RJM, DEW-M, AH, NA, PI, EB, NC, JK, JLC served as Pre-Vent site PI and supervised the studies that generated the data used for this analysis and manuscript. All authors reviewed and approved the final version of the manuscript. Authors listed as Pre-Vent Investigators contributed to data curation or project administration.

## Notes

### Competing Interest Statement

Some authors have financial conflicts of interest. JRM and DEL own stock in Medical Prediction Sciences Corporation. JRM is a consultant for Nihon Kohden Digital Health Solutions, proceeds donated to the University of Virginia. ZAV is a consultant for Medtronic. All other authors have no financial conflicts to disclose. No authors have any non-financial conflicts of interest to disclose.

### Funding Statement

We acknowledge the following NIH grants for funding the work presented in this manuscript:
University of Virginia (NCT03174301): U01 HL133708, K23 HD097254, HL133708-05S1
Case Western Reserve University: U01 HL133643
Northwestern University: U01 HL133704
University of Alabama at Birmingham: U01 HL133536, K23 HL157618
University of Miami: U01 HL133689
Washington University: U01 HL133700, F

### Author Declarations

Pre-Vent (ClinicalTrials.gov identifier NCT03174301). Institutional Review Boards (IRBs) reviewed the study and approved collection of clinical and cardiorespiratory monitoring data at all participating centers with a waiver of consent at three centers and informed consent required at two centers. Approval was obtained from the Hospital Institutional Review Board for each study. University Hospitals Cleveland Medial Center IRB for Human Investigation: Approval number: 04-16-26 Approval date: 7/19/2017 Study title: The Association between Intermittent Hypoxia and Later Respiratory Morbidity (Pre-Vent Study) Ann & Robert H. Lurie Children's Hospital of Chicago IRB: Approval number: 2017-1076 Approval date: 4/11/2017 Study title: Prematurity-Related Ventilatory Control (Pre-Vent): Role in Respiratory Outcomes: Multicenter Common Protocol The University of Alabama at Birmingham IRB for Human Use: Approval number: 170428003 Approval date: 6/2/2017 Study title: Prematurity-Related Ventilatory Control (Pre Vent): Role in Respiratory Outcomes: Multicenter Common Protocol University of Miami IRB: Approval number: 20170402 Approval date: 6/19/2017 Study title: Prematurity-Related Ventilatory Control (Pre-Vent): Role in Respiratory Outcomes - Multicenter Common Protocol Washington University IRB: Approval number: 201611138 Approval date: 10/31/2017 Study title: Physiologic Biomarkers Predicting Ventilatory Instability and Hypoxemia in Premature Infants University of Virginia IRB for Health Sciences Research: Approval number: 19606 Approval date: 3/1/2017 Study title: Prematurity-Related Ventilatory Control (Pre-Vent): Role in Respiratory Outcomes - Multicenter Common Protocol Procedures were followed in accordance with the ethical standards of the responsible committee on human experimentation and with the Helsinki Declaration of 1975. Oversight was also provided by an observational and safety monitoring board, appointed by the NHLBI.

