## Supplementary appendix for "Apnea, Intermittent Hypoxemia, and Bradycardia Events Predict Late-Onset Sepsis in Extremely Preterm Infants"

**Table of Contents**

Page 3: Pre-Vent Investigators

Page 5: Supplementary methods- Cardiorespiratory metrics

Page 5 - 6: Supplementary methods- Institutional Review Board Approval Information

Page 7: Table S1. Number of infants by gestation age.

Page 8: Table S2. Patient-days with cardiorespiratory monitoring data available for feature calculation.

Page 9: Table S3 Univariate AUCs of clinical variables to predict sepsis.

Page 10. Figure S1. Distribution of sepsis events by age

Page 11. Figure S2. Calibration of the clinical model + 4 physiologic variables + ventilator status.

**Pre-Vent Investigators:**

The following investigators, in addition to those listed as authors, participated in this study:

Pre-Vent Investigators: Katy N. Krahn and Amanda M. Zimmet, University of Virginia Center for Advanced Medical Analytics, Charlottesville, VA; Bradley S Hopkins, Erin K Lonergan, Casey M. Rand: Ann & Robert H. Lurie Children’s Hospital of Chicago, and Pediatric Autonomic Medicine, Stanley Manne Children’s Research Institute, Chicago, IL; Arlene Zadell, Department of Pediatrics, University Hospitals Cleveland Medical Center, Rainbow Babies and Children’s Hospital, Cleveland, OH; Arie Nakhmani, Department of Electrical and Computer Engineering, University of Alabama at Birmingham, Birmingham, AL; Waldemar A. Carlo, Deborah Laney, and Colm P. Travers, Division of Neonatology, Department of Pediatrics, University of Alabama at Birmingham School of Medicine, Birmingham, AL; Silvia Vanbuskirk, Carmen D’Ugard, Ana Cecilia Aguilar and Alini Schott, Division of Neonatology, Department of Pediatrics, University of Miami Miller School of Medicine, Holtz Children’s Hospital – University of Miami/Jackson Memorial Medical Center, Miami, FL; and Julie Hoffmann and Laura Linneman, Washington University School of Medicine in St. Louis, St. Louis, MO.

NIH/NHLBI

Neil Aggarwal, MD: National Institutes of Health, National Heart, Lung and Blood Institute, Division of Lung Diseases, Bethesda, MD

Lawrence Baizer, PhD: National Institutes of Health, National Heart, Lung and Blood Institute, Division of Lung Diseases, Bethesda, MD

Peyvand Ghofrani, MDE: National Institutes of Health, National Heart, Lung and Blood Institute, Division of Lung Diseases, Bethesda, MD

Aaron D. Laposky, PhD: National Institutes of Health, National Heart, Lung and Blood Institute, Division of Lung Diseases, Bethesda, MD

Aruna Natarajan MD, PhD: National Institutes of Health, National Heart, Lung and Blood Institute, Division of Lung Diseases, Bethesda, MD

Barry Schmetter, BS: National Institutes of Health, National Heart, Lung and Blood Institute, Division of Lung Diseases, Bethesda, MD

OSMB

Estelle B. Gauda, MD (Chair): University of Toronto Hospital for Sick Children, Division of Neonatology, Toronto, Ontario

Jonathan M. Davis, MD: Tufts Clinical and Translational Science Institute, Division of Newborn Medicine, Boston, MA

Roberta L. Keller, MD: University of California, San Francisco School of Medicine, Department of Pediatrics, San Francisco CA

Robinder G. Khemani, MD: Children’s Hospital Los Angeles, Department of Anesthesiology and Critical Care Medicine, Los Angeles, CA

Renee H. Moore, PhD: Drexel University Department of Epidemiology and Biostatistics, Philadelphia, PA

Elliott M. Weiss, MD, MSME: Seattle Children's Hospital, Department of Pediatrics, Division of Bioethics, Seattle, WA

Washington University: U01 HL133700, K23 NS111086

**Supplementary Methods:**

**Cardiorespiratory metrics justification:**

In the Pre-Vent study, each of the 5 physiologic features (apnea, periodic breathing, IH80, IH90, and bradycardia) were summarized daily using three different measures: (1) Counts of events measured in number per day (frequency), (2) exposure, or total event time measured in minutes per day, and (3) duration per event (DPE) measured in seconds per event. These measures are usually highly correlated, therefore, one measure per feature was chosen for analysis, for a total of five features. In the primary analysis of the Pre-Vent study, the duration per IH80 and IH90 events were shown to be the best univariate feature for predicting unfavorable respiratory outcomes [(21)](https://sciwheel.com/work/citation?ids=14900131&pre=&suf=&sa=0) and therefore were chosen for this analysis. We chose counts of apnea and bradycardia events and exposure to periodic breathing per day based on additional prior studies [(9, 24)](https://sciwheel.com/work/citation?ids=6748704,5426005&pre=&pre=&suf=&suf=&sa=0,0).

**Institutional Review Board Approval Information:**

Approval was obtained from the Hospital Institutional Review Board for each study.

University Hospitals Cleveland Medial Center IRB for Human Investigation:

Approval number: 04-16-26

Approval date: 7/19/2017

Study title: The Association between Intermittent Hypoxia and Later Respiratory Morbidity (Pre-Vent Study)

Ann & Robert H. Lurie Children's Hospital of Chicago IRB:

Approval number: 2017-1076

Approval date: 4/11/2017

Study title: Prematurity-Related Ventilatory Control (Pre-Vent): Role in Respiratory Outcomes: Multicenter Common Protocol

The University of Alabama at Birmingham IRB for Human Use:

Approval number: 170428003

Approval date: 6/2/2017

Study title: Prematurity-Related Ventilatory Control (Pre Vent): Role in Respiratory Outcomes: Multicenter Common Protocol

University of Miami IRB:

Approval number: 20170402

Approval date: 6/19/2017

Study title: Prematurity-Related Ventilatory Control (Pre-Vent): Role in Respiratory Outcomes - Multicenter Common Protocol

Washington University IRB:

Approval number: 201611138

Approval date: 10/31/2017

Study title: Physiologic Biomarkers Predicting Ventilatory Instability and Hypoxemia in Premature Infants

University of Virginia IRB for Health Sciences Research:

Approval number: 19606

Approval date: 3/1/2017

Study title: Prematurity-Related Ventilatory Control (Pre-Vent): Role in Respiratory Outcomes - Multicenter Common Protocol

Procedures were followed in accordance with the ethical standards of the responsible committee on human experimentation and with the Helsinki Declaration of 1975.

| **Gestational Age (weeks)** | **N (%)** |
| --- | --- |
| 22 | 12 (1.6%) |
| 23 | 70 (9.5%) |
| 24 | 68 (9.2%) |
| 25 | 137 (18.6%) |
| 26 | 131 (17.8%) |
| 27 | 131 (17.8%) |
| 28 | 188 (25.5%) |

**Table S1. Number of infants by gestation age.** Values are presented as number and percentage.

| **Cardiorespiratory Feature** | **All times**  *With monitoring data* | **Off ventilator times**  *With monitoring data* | **On ventilator times**  *With monitoring data* |
| --- | --- | --- | --- |
| Bradycardia | 48,192 | 35,285 | 12,907 |
| IH80 | 47,806 | 35,127 | 12,679 |
| IH90 | 47,806 | 35,127 | 12,679 |
| Apnea |  | 35,391 |  |
| Periodic Breathing |  | 35,322 |  |

**Table S2.** **Patient-days with cardiorespiratory monitoring data available for feature calculation.** Patient-days varied according to the availability of the monitoring data used to calculate each of the five cardiorespiratory features. Bradycardia calculation used ECG-derived heart rate, intermittent hypoxia < 80% (IH80) and <90% (IH90) calculation used pulse-oximeter-derived SpO_2_, and apnea and periodic breathing metrics used chest-impedance waveforms.

| **Variable** | **AUC** | | |
| --- | --- | --- | --- |
|  | **All times** | **On ventilator times only** | **Off ventilator times only** |
| Sex | 0.507 | 0.490 | 0.503 |
| Small for GA* | 0.543 | 0.518 | 0.503 |
| FiO_2_ High* | 0.609 | 0.495 | 0.554 |
| Supplemental O_2_* | 0.629 | 0.510 | 0.591 |
| GA* | 0.639 | 0.553 | 0.553 |
| FiO_2_* | 0.661 | 0.509 | 0.590 |
| Positive Pressure* | 0.675 | 0.500 | 0.644 |
| Vent Status* | 0.689 | 0.500 | 0.500 |
| PMA* | 0.722 | 0.600 | 0.701 |
| Respiratory Support* | 0.729 | 0.500 | 0.644 |

**Table S3. Univariate AUCs of clinical variables to predict sepsis.** Positive pressure is defined as a 2-level variable, (1) no positive pressure, (2) positive pressure or invasive mechanical ventilation. Respiratory support is defined as a 3-level variable, (1) no respiratory support, (2) positive pressure, and (3) invasive mechanical ventilation. High FiO_2_ is defined as greater than 30%. *p<0.05 on the logistic regression coefficient adjusted with the robust covariance matrix. Abbreviations: PMA = post menstrual age, GA = gestational age, FiO_2_ = fraction of inspired oxygen

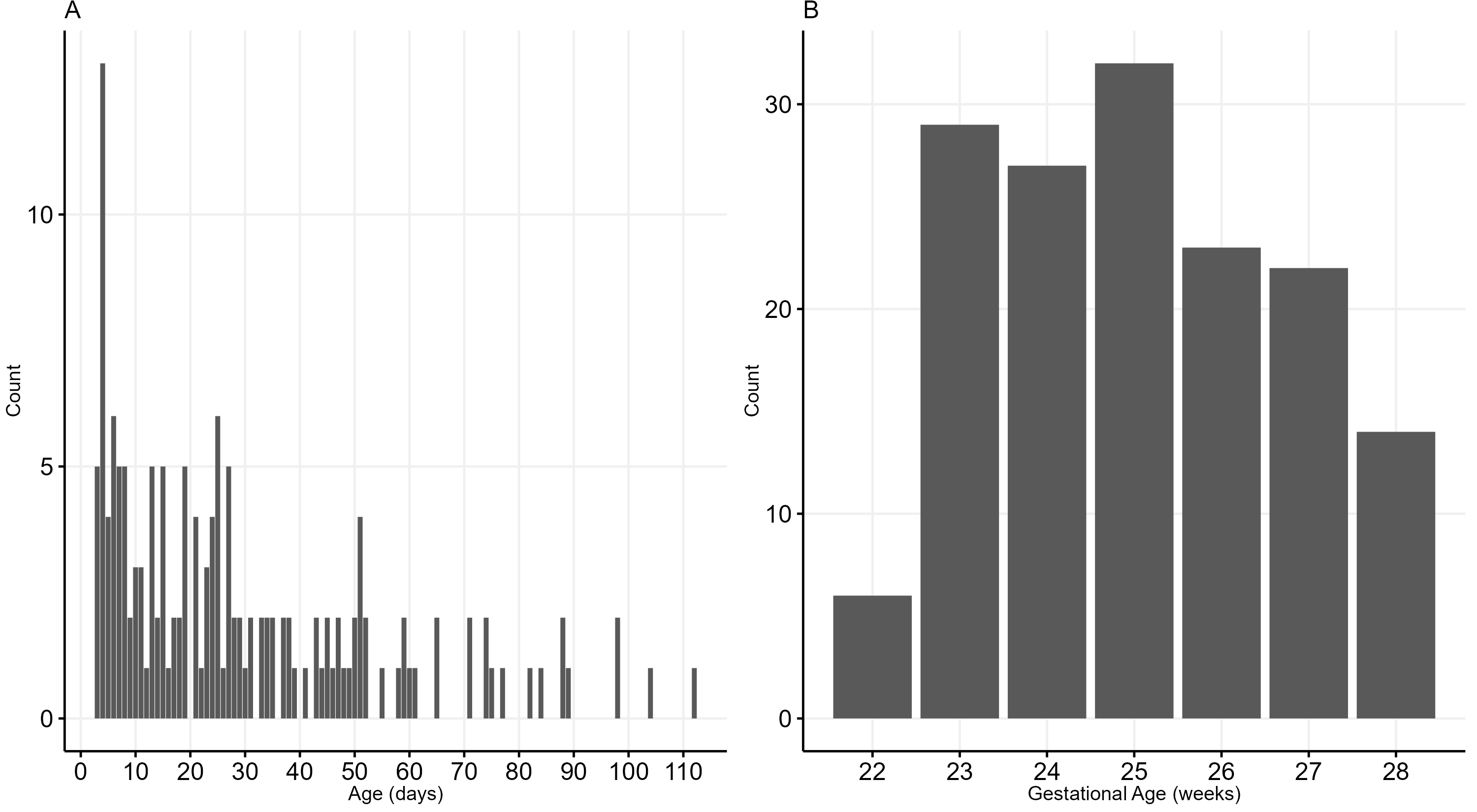

**Figure S1. Distribution of sepsis events by age**. (A) Bar plot of sepsis events by age in days. (B) Bar plot of sepsis event by gestation age.

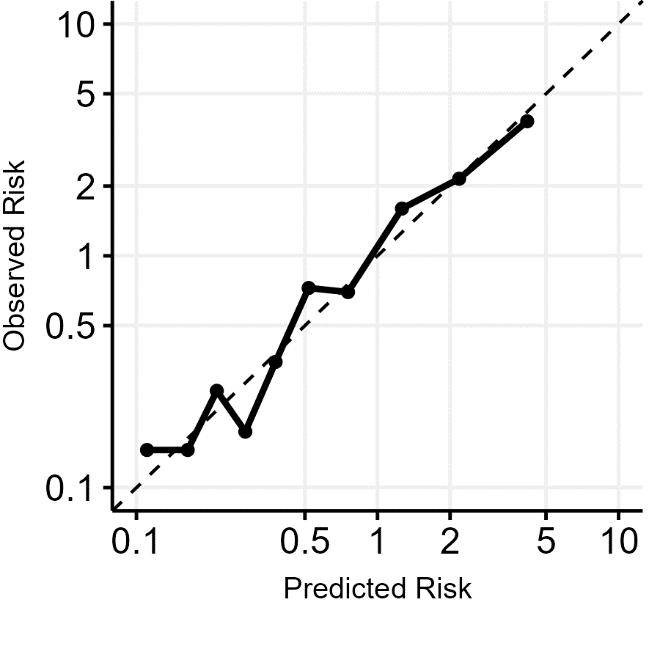

**Figure S2. Calibration of the clinical model + 4 physiologic variables + ventilator status.** Predicted risk relative to average is on the abscissa and observed risk relative to average is on the ordinate. Each point represents one decile of predicted risk. The line of identity is shown as a dashed line.
